# The Acute Effects of Physical Exercise Breaks on Cognitive Function During Prolonged Sitting: A First Quantitative Evidence

**DOI:** 10.1101/2022.01.29.22270085

**Authors:** Jinming Li, Fabian Herold, Sebastian Ludyga, Qian Yu, Xingyu Zhang, Liye Zou

## Abstract

**Purpose:** This systematic review and meta-analysis aims to investigate the cognitive benefits of breaking up prolonged sitting (PS) by acute physical exercises (PE).

**Methods:** A systematic literature search was performed in six electronic databases (PubMed, Scopus, Web of Science, PsycINFO, SPORTDiscus, and Cochrane Library) to identify cross-over studies with a pre-posttest design that examined the effects of PE breaks during 3 to 5 h of prolonged sitting on cognitive performance. A meta-analysis was performed using a random effects model, with subgroup analyses investigating dose-response effects and differences between cognitive domains. Additionally, study quality was rated using the PEDro scale.

**Results:** Thirteen randomized controlled trials (RCTs) with a total of 295 participants (171 female and 124 male) were included in this systematic review. Thereof, nine studies were included in our meta-analysis. We observed that during PS, PE breaks did not improve overall cognitive performance (Hedges’ g=-0.043[-0.158,0.073]). There was no between-study heterogeneity indicated. This is further supported by the subgroup analyses showing no differences in effect sizes between cognitive domains and different exercise intensities.

**Conclusions:** Our findings suggest that interrupting PS by PE breaks neither improved, nor impaired cognitive performance. Consequently, PE breaks during PS allows to integrate regular physical activity in daily routines (i.e., through PE breaks) without compromising the performance of cognitively demanding tasks.

## Introduction

In the literature it has been conjectured that physical inactivity will become the biggest public health problem of the 21^st^ century [1]. In this context, physical inactivity is characterized as the failure to comply with international physical activity recommendations [2]. Although conceptually different from physical inactivity, the increase of sedentary behavior (SB) which refers to sitting (e.g., prolonged sitting being characterized by sitting periods > 30 minutes [3-5]), reclining, or lying posture with an energy expenditure of ≤1.5 metabolic equivalents [6], further underlines the need to intervene. There is evidence that the prevalence of SB among adults has reached 69% in the USA [7], and that 60.9% of younger adults such as Peruvian university students under 20 years old sitting more than 8 hours per day [8]. In addition, Chinese and American college students spent roughly 10 and 15 hours with SB per day, respectively [9, 10]. Such unhealthy lifestyle behaviors have been linked to the greater probability of developing a variety of chronic diseases (e.g., hypertension, obesity) [11] and even pre-mortality [12].

Recently, it has been recognized that high levels of SB (e.g., watching TV) in adults can have detrimental effects on cognitive performance [13, 14] although this finding is not universal [15, 16]. In this venture, it has been observed that in adults, an increased level of SB is associated with worse performance in executive functioning [17] and in verbal memory [18]. Recalling that high levels of SB might negatively influence cognitive performance and the evidence indicated that individuals spent almost 60% of their everyday waking hours in SB [19], there is a need to counteract its detrimental health consequences. Especially acute and chronic physical exercises (defined as specific, structured, and planned forms of physical activity [20-23]) seems to be a valuable option to do so, given the evidence that acute [24-27] and chronic physical exercises [28-32] can improve cognitive performance regardless of age [33] and environmental condition [34].

Recently health professionals and researchers advocated that interrupting prolonged sitting PS by physical exercise (PE) breaks may be an effective approach to render positive health outcomes (e.g., cognitive performance) [35] as breaking up PS (e.g., while studying or working) by PE breaks seems to be a practical and feasible way to integrate acute exercises in our daily routines. Indeed, a cross-over study (N = 11 Qatar females) investigated the effects of interrupting 5-horus of sitting by 3-min walking every 30 minutes on cognitive performance, suggesting that PE breaks improved reaction time (as measured by the Stroop task) as compared to PS (i.e., sitting for 5 hours) [36]. In a comparable manner, Mullane et al [37] observed in younger, healthy adults that PE breaks(i.e., standing, walking, and cycling) performed during PS (i.e., sitting for a period of 8 hours) leads to significantly better executive functioning (as measured by the Set Shifting Test) and working memory performance (as measured by 1-Back test) in walking and cycling conditions, but not in standing condition, as compared to PS. However, Sperrlich et al. recruited 12 healthy, young students and ask them to engage in two different conditions (i.e., being seated for 3 hours or sitting for the same period while interrupted PS with a 6-min exercise after 1 hour), but only noticed a non-significant difference in the Stroop task between the two conditions [38]. Such mixed results across different studies may not allow to draw a clear conclusion regarding the effects of PA break during PS and might point towards specific dose-response relationships.

While some previous reviews and meta-analysis focused on the effects of acute exercises on cognitive performance [24-27], up to now, there are no meta-analytic reviews investigating the effects of interrupting PS by PE breaks on measures of cognitive performance. With regard to effects of PE breaks during PS on cognitive performance, there is only a narrative review that highlights physiological mechanisms by which PE breaks may influence cognitive performance during prolonged sitting [39], However, a quantitive synthesis that allows the estimation of the effect is lacking.

To address this gap in literature, the current systematic and meta-analysis is devoted to answer the following main questions: (i) Does breaking up PS by acute PE breaks lead to improvements concerning measures of cognitive performance? and (ii) Does specific exercise characteristics of the PE breaks (e.g., type of physical exercise, exercise intensity, and/or exercise duration) moderate the effect of PE breaks during PS on cognitive performance? To answer these two major research questions, this systematic review and meta-analysis searched six electronic databases and analyzed the data of 295 participants who were enrolled in 13 randomized controlled trials with crossover design.

## Methods

### Protocol and registration

According to the checklist of Preferred Reporting Items for Systematic Reviews and Meta-Analyses (PRISMA) are search protocol was developed [40], and this research was prospectively registered in the International Prospective Register of Systematic Reviews (PROSPERO, CRD42021224949).

### Search strategy and process

Two independent reviews conducted a systematic literature search in December 2020 adhering to the established guidelines [41] and if necessary, a third researcher was consulted. The following six electronic databases were used for the systematic literature search: PubMed, Scopus, Web of Science, PsycINFO, SportDiscus, and Cochrane Library. We limited the search to title and abstract and used the following search terms: “prolonged sitting” OR “uninterrupted sitting” OR “interrupted sitting” OR “acute exercis*” OR “physical activity break*” OR “walk*” OR “cycling” AND “cogniti*”.

### Inclusion and exclusion criteria

We used the established PICOS principle to screen for relevant articles. “PICOS” represents the following categories: participants (P), intervention (I), comparison s (C), outcomes (O), and study design (S) [40, 42].

In the present meta-analysis study, the following inclusion and exclusion criteria were applied: (P) we used no restrictions regarding age groups and pathologies; (I) we include studies if they investigated the effect of acute physical activity breaks within or compared to a single bout of prolonged sitting on cognitive functions. In this context, we refer to “prolonged sitting” as sitting time > 30 minutes[3-5]. However, to avoid an overlap with existing reviews (e.g., investigating the influence of acute exercises[24, 27] and given that in ecologically valid settings (e.g., college classes, working in an office) sitting times are normally longer (e.g., 45 minutes in college classes), we intentionally included only studies which investigated on overall sitting time ≥ 2h (e.g., 4x bouts of 30 minutes of seated rest interrupted by short physical activity breaks); (C) we do not include studies which solely compare a bout of acute physical exercise (e.g., cycling for 45 minutes) to a rest condition (e.g., sitting for 45 minutes), but we included studies which incorporated short physical activity breaks within a prolonged sitting period (e.g., physical activity breaks of a total duration of ≤ 15 minutes within/after a prolonged sitting bout of ≥ 30 minutes). Furthermore, we do not consider studies as relevant if they interrupt prolonged sitting only by standing; (O) we considered studies as relevant if the assessed changes in cognitive performance (e.g., using standardized cognitive tests such as Stroop test to assess cognitive performance) but excluded studies which solely assessed biological changes without investigating a relationship to cognitive performance (e.g., changes in cerebral blood flow without assessing cognitive performance); (S) we included only acute interventional studies in this meta-analysis. In addition, we exclude studies if they were not written in English and have not been published in a peer-reviewed journal.

### Data extraction

We extracted the following information from the relevant studies: (i) first author and year of publication, (ii) population characteristics including age, gender, cognitive status, (iii) characteristics regarding the intervention and comparison condition (e.g., study design; duration of prolonged sitting, activities allowed during prolonged sitting), (iv) cognitive testing (e.g., assessed cognitive domain), and (vi) outcomes (e.g., changes in cognitive performance in response to prolonged sitting). Of note, cognitive measures were categorized into attention, executive function, and memory according to previous reviews ([24, 32, 34]). In addition, exercise intensity was reported in 12 studies, while calf raising was determined (by our research team) as the light exercise intensity.

### Quality Assessment

The quality of each study was assessed using the Physiotherapy Evidence Database (PEDro) scale which consists of 10 items (10–9 = excellent, 8–6 = good, 5–4 = fair, < 4 = poor[43]), separately by 2 review authors. The assessment was conducted based on the following categories: randomization, concealed allocation, baseline equivalence, blinding of participants, assessors, and instructors, the retention rate of ≥85%, missing data management (intent-to-treat analysis), between-group analysis, and point measure and measures of variability. For each category a point was given when they conformed to the standard of scale.

### Statistical Analyses

For outcomes of interest, mean and SD values of post-minus-baseline difference, and sample size were extracted for both PE break and control groups/condition. In those studies where authors did not report SD, standard error and 95% confidence interval were used to calculate SD in light of the recommended formula: (1) SD=SE* SQRT (N), where N is the sample size; (2) SD=SQRT (N) * [(UCI − LCI)/3.92], where U= upper CI, L= lower CI. For studies, which provided graphical results, the aforementioned quantitative data were obtained through ImageJ (V.1.50i, https://imagej.nih.gov/ij/): the length of the axes as well as the length of histogram were measured and then calibrated. If the relevant information could not be obtained through the aforementioned methods, we directly requested data from the corresponding authors of the respective studies. When authors reported cognitive assessments over multiple time points, the change between the initial assessment and the assessment closest to 5 hours was used for quantitative synthesis. In case multiple tasks were used to assess a single cognitive domain, the effect sizes were pooled to form a single outcome. Additionally, either reaction time or accuracy was used to calculate effect sizes, because using both measures can bias the estimation of the true effect. The selection was based on the primary outcome of the cognitive task.

To obtain the magnitude of PE break intervention effects on cognitive function, Hedges’ g with 95% confidence interval (95% CIs) was calculated and its value can be interpreted as small (0.2), moderate (0.5), and large (0.8). Pairwise meta-analyses for cognitive function and its components were performed based on the random-effects model, while the heterogeneity of between-study comparisons was assessed using the I^2^ statistic and its value can be interpreted as low (25%), medium (50%), and large (75%). Publication bias was assessed using the Egger’s test along with a funnel plot. In case heterogeneity was detected, subgroup analyses on cognitive domain (executive function, attention, memory) and exercise intensity (light, moderate, high) were performed.

## Results

### Study selection

43862 articles were identified through electronic and manual search. After removing duplicates, 31550 articles remained, and a screening of titles and abstracts was performed. After the title and abstract screening, 21 articles were considered as potentially relevant, of which eight articles were deemed ineligible and being excluded after full text screening. Finally, 13 articles met our inclusion criteria, but only 9 studies were included for meta-analytical data analysis because four studies did not report usable data. Given the fact that this is a newly emerging research direction, the four studies were included in the systematic review (i.e., descriptive analysis) but were not considered in the meta-analytical data analysis. Figure 1 displays a flowchart of the article retrieval process.

**Figure 1.**
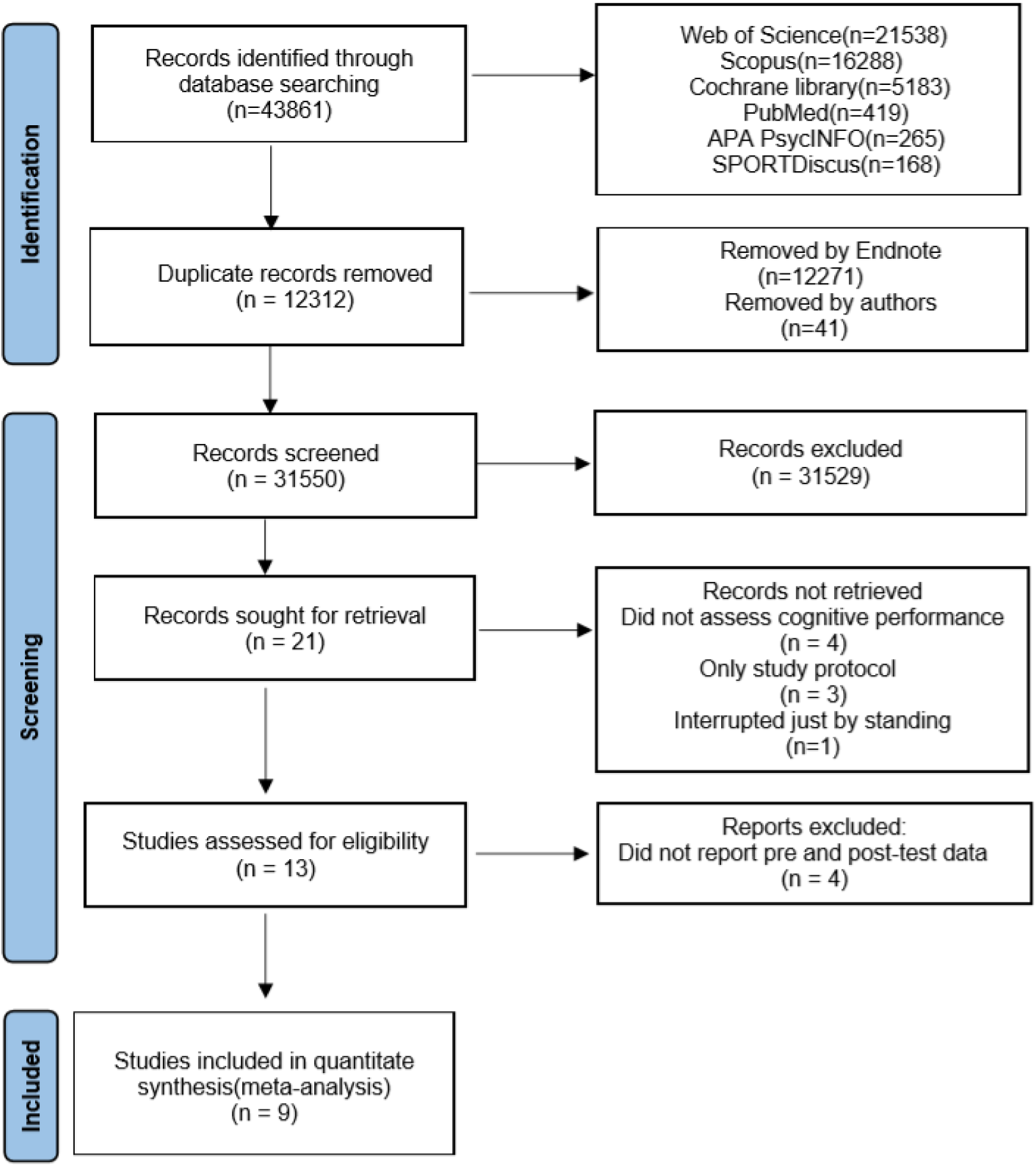
Flowchart of the article retrieval process

**Figure 2.**
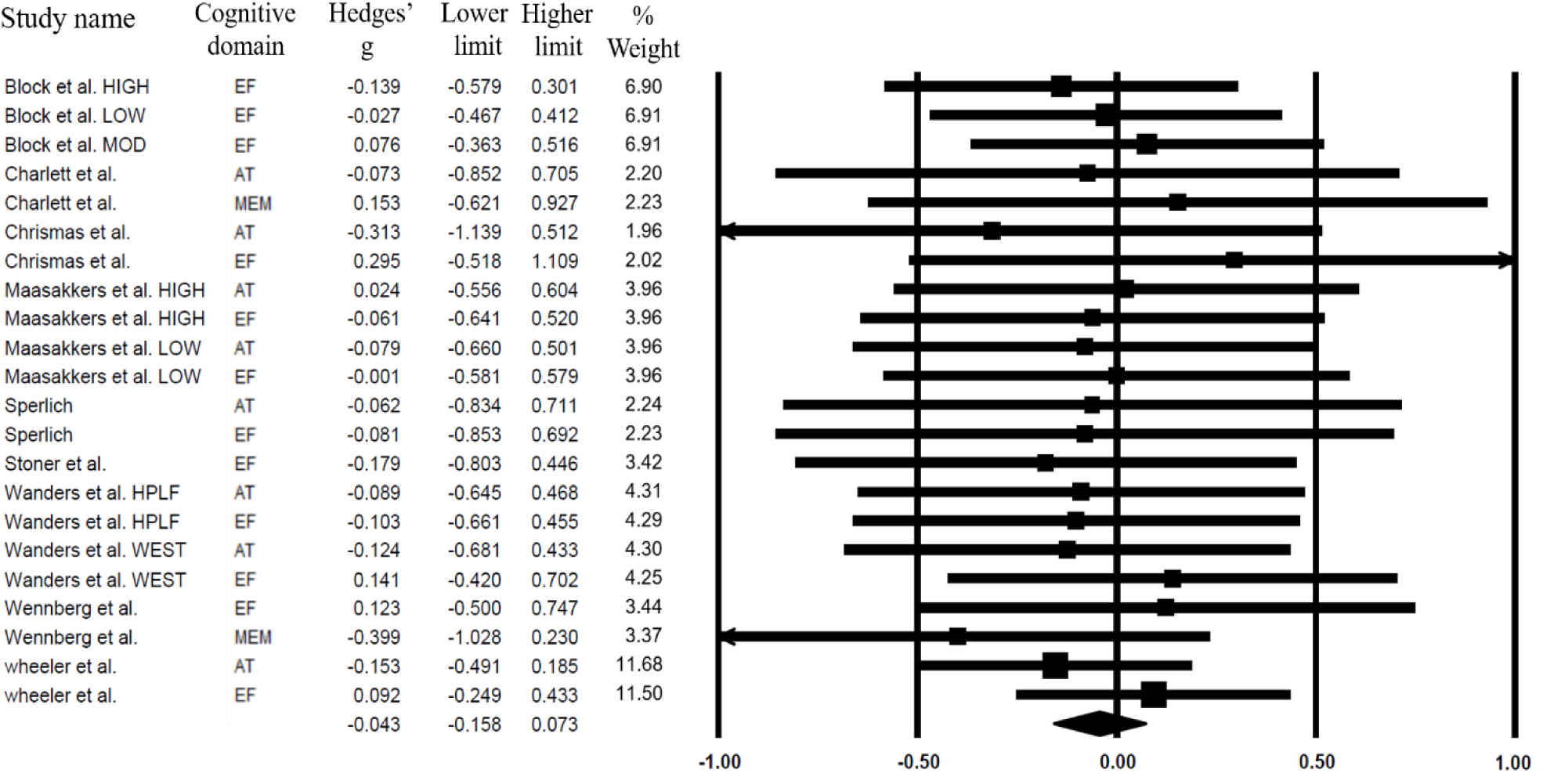
Overall effect of PE breaks on cognition

**Figure 3.**
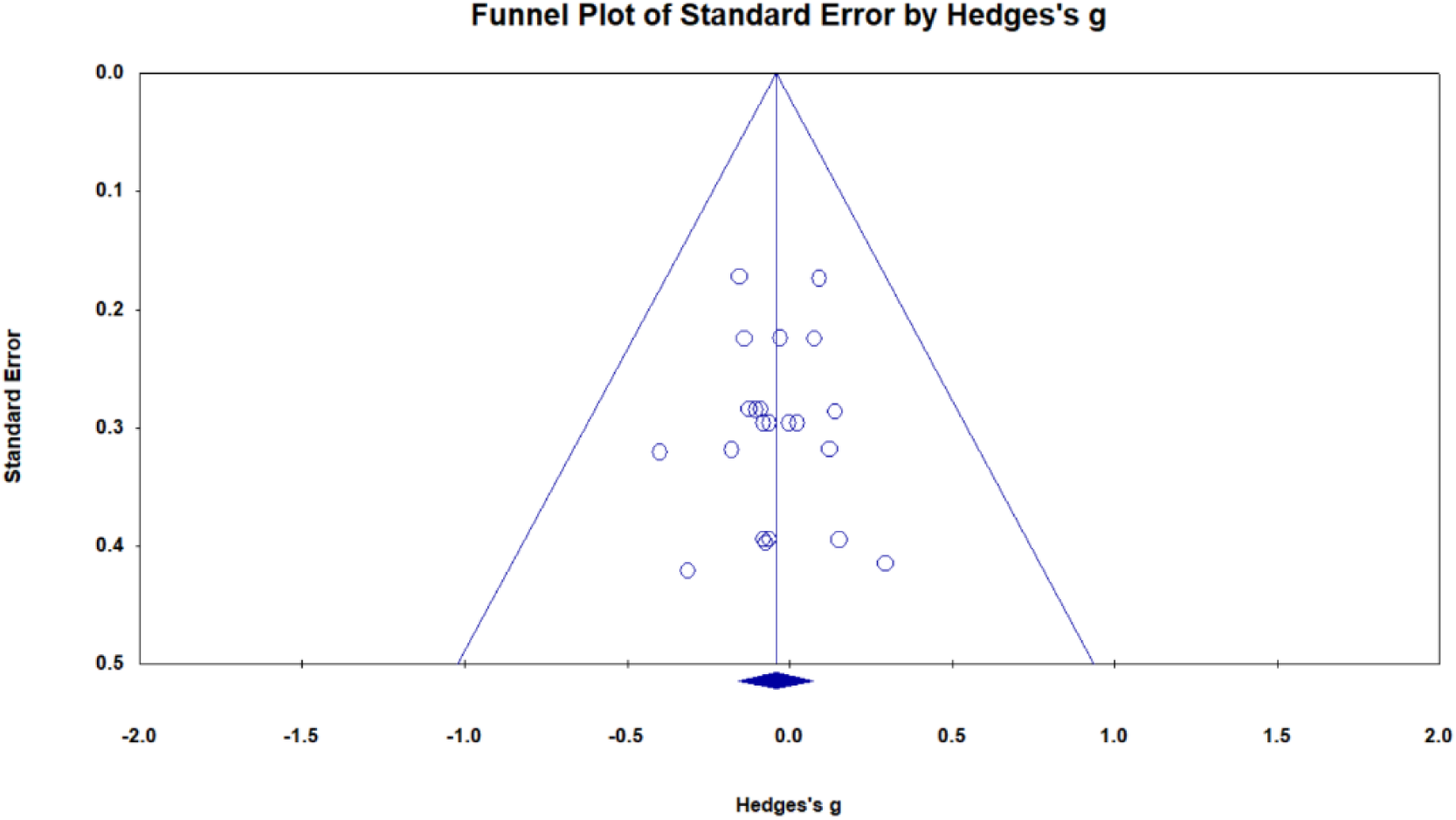
Funnel Plot

### Characteristics of Included Studies

We included thirteen randomized cross-over studies that have been conducted in six different countries (American, Netherlands, Australia, German, Qatar and Britain) and were published between 2016 to 2020. A total of 295 participants (171 female and 124 male) were included in this review, with sample size of each individual study ranging from 6 to 67; the mean age ranged from 9 to 78 years. The majority of the studies investigated younger adults (aged 22-30)[36-38, 44-47] and older adults (aged 60-78)[16, 48-51] and only one study focused on children (aged 9 years old on average)[52]. Remarkably, in seven studies overweight and/or obese individuals (BMI ≥ 25 kg/m^2^) were studied[16, 37, 46, 48-51].

With respect to exercise characteristics of the PE break, the following types of physical exercises were used: (i) walking [16, 36, 37, 44, 47, 49-51], (ii) cycling [37, 49], (iii) standing[37], (iv) bodyweight resistance exercise [45] including calf raises [46], and (v) intermittent exercise [38, 52]. Different exercise intensity (light, moderate, and/or high) were used in insolation or conjunctively across 13 studies (see Table 1 for a detailed overview). In the included studies, the exercise duration of PE breaks lasted from 30 seconds to 30 minutes (see Table 1 for a detailed overview). Of note, participants were allowed to engage in several activities (e.g., TV watching, reading, and working on a laptop) during the PS period. The study characteristics are shown in more detail in Table 1.

**Table 1.** Summary of characteristics of all studies meeting the inclusion criteria

| Study | Study Design | Participants | Prolonged Sitting (duration) | Physical activity break (type, length, intensity, duration) | Outcome of Interest (Instrument) |
| --- | --- | --- | --- | --- | --- |
| Bergouignan et al., 2016 | Crossover RCT: 3 conditions | N = 30 (21 female/9 male)<br>Age: $30 \pm 5.6$ years<br>BMI: $23 \pm 2.4$ kg/m<sup>2</sup><br>Cognitive status: healthy | C1: 6-hour sitting | C2: 6-hour sitting (only 30min moderate-intensity walk at the beginning)<br>C3: 6-hour sitting (interrupted sitting per 60min with 5min moderate-intensity walk)<br>Duration (C3): 5min/bout x 6 bouts = 30min | Attention (Comprehensive Trail Making Test)<br>Executive function (Modified Eriksen Flanker Task) |
| Mullane et al., 2016 | Crossover RCT: 4 conditions | N = 9 (7 female/2 male)<br>Age: $30 \pm 15$ years<br>BMI: $29 \pm 3.0$ kg/m<sup>2</sup> (overweight)<br>Cognitive status: healthy | C1: 6-hour sitting | C2 (standing), C3 (cycling), & C4 (walking):<br>6-hour sitting (interrupted sitting with 10min break at 08:50 & 09:50h, 15min break at 10:45 & 11:45h, 20min break at 12:40 & 13:20h, and 30min break at 14:00 & 15:30h)<br>Duration: 150min of low intensity standing, cycling, walking, respectively | Attention (Detection Test)<br>Executive function (Set-Shifting Test, One Back Test) |
| Wennberg et al., 2016 | crossover RCT: 2 conditions | N=19 (9 female/10 male)<br>Age: $59.7 \pm 8.1$ years<br>BMI: $31.5 \pm 4.7$ kg/m<sup>2</sup> (overweight/obese)<br>Cognitive status: healthy | C1: 7-hour sitting | C2: 7-hour sitting (150min sitting and then interrupted sitting per 30 min with 3min light-intensity walk on a motorized treadmill).<br>Duration: 3min/bout * 10 bouts = 30min | Memory (Face-Name Association Test)<br>Executive function (n-back, letter memory, Eriksen flanker task and modified Stroop color-word task) |
| Duvivier et al., 2017 | Crossover RCT: 2 conditions | N = 24 (14 female/13 male)<br>Age: $64 \pm 7$ years<br>BMI: $29 \pm 2$ kg/m<sup>2</sup> (overweight/obese)<br>Cognitive status: healthy | C1: sitting 13.5h/day | C2: sitting 7.6h/day, standing 4.0h/day, light-intensity walking 4.3 h/day | Memory (Rey's Verbal Learning Test)<br>Executive function (Trail Making Test)<br>Attention (Attention Network Test) |
| Sperlich et al., 2018 | Crossover RCT: 2 conditions | N=12 (7 female/5 male)<br>Age: $22 \pm 2$ years<br>BMI: $21.7 \pm 2.1$ kg/m<sup>2</sup><br>Cognitive status: healthy | C1: 3-hour sitting | C2: 3-hour sitting (60min sitting and then 6min high-intensity interval training)<br>Duration: 6min | Executive function (Stroop Test) |
| Block et al., 2018 | Crossover RCT: 4 conditions | N=39 (21 female/18 male)<br>Age: $9.0 \pm 0.2$<br>BMI: $18.5 \pm 0.6$ kg/m<sup>2</sup><br>Cognitive status: healthy | C1: 8-hour sitting | C2, C3 & C4: 8-hour sitting (interrupted sitting per 20min with 2min low-, moderate-, and high-intensity exercise)<br>Duration : 2min/bout x 20 bouts = 40min | Executive function (40-digit Addition & Subtraction) |
| Vincent et al., 2018 | Crossover RCT: 2 conditions | N = 6 (6 male)<br>Age: $27 \pm 3.7$ years<br>BMI: $24.84$ kg/m<sup>2</sup><br>Cognitive status: healthy | C1: 8-hour sitting | C2: 8-hour sitting (interrupted sitting per 30min with 3min light-intensity walk)<br>Duration: 3min/bout x 17 bouts = 51min | Attention (Psychomotor vigilance test and Digit Symbol Substitution Test) |
| Stoner et al., 2019 | Crossover RCT: 2 conditions | N = 20 (14 female/6 male)<br>Age: 21.7 ± 5.2 years<br>BMI: 25.5 ± 6.1 kg/m<sup>2</sup> (overweight)<br>Cognitive status: healthy | C1: 3-hour sitting | C2: 3-hour sitting (interrupted sitting at Min 15, Min 20, & then per 10min till Min 170, with 30-second calf raises)<br>Duration: 30 seconds/bout x 17 bouts = 8.5min | Executive function (Stroop Test) |
| Christmas et al., 2019 | Crossover RCT: 2 conditions | N = 11 (females)<br>Age: 29.19 ± NR years<br>BMI: 22.72 kg/m<sup>2</sup><br>Cognitive status: healthy | C1: 5-hour sitting | C2: 5-hour sitting (Interrupted sitting per 30min with 3min moderate-intensity walk on a motorized treadmill)<br>Duration: 3min/bout x 9 bouts = 27min | Executive function (Stroop Task, Serial 3 & 7 subtractions Task)<br>Attention (Simple reaction time, Choice reaction time and Rapid visual information processing.) |
| Wheeler et al., 2019 | Crossover RCT: 3 conditions | N = 67 (35 female/32 male)<br>Age: 67 ± 7 years<br>BMI: 31.2 ± 4.1 kg/m<sup>2</sup> (obese)<br>Cognitive status: healthy | C1: 8-hour sitting | C2: 8-hour sitting (60min sitting & then 30min moderate-intensity walk)<br>C3: 8-hour sitting (60min sitting & then 30min moderate-intensity walk, followed by per 30min with 3min light-intensity walk)<br>Duration(C3): 30min+3min/bout x 13 bouts = 69min | Executive function (Groton Maze Learning Test, One Back Test, Two Back Test and One Card Learning Test)<br>Attention (Detection Test and Identification Test) |
| Maasakkers et al., 2020 | Crossover RCT: 4 conditions | N = 22 (9 female/13 male)<br>Age: 78 ± 5.3 years<br>BMI: 26 ± 4.0 kg/m<sup>2</sup> (overweight)<br>Cognitive status: healthy | C1 (high mental activity)<br>C2 (low mental activity): 3-hour sitting | C3 (high mental activity)<br>C4 (low mental activity): 3-hour sitting (Interrupted sitting per 30min with 2-min light-intensity walk).<br>Duration: 2min/bout * 5 bouts = 10min | Attention and Executive function (The Test of Attentional Performance Battery) |
| Charlett et al., 2020 | Crossover RCT: 2 conditions | N = 12 (7 female/5 male)<br>Age: 25 ± 6 years<br>BMI: 24.7 ± 4.9 kg/m<sup>2</sup><br>Cognitive status: healthy | C1: 5-hour sitting | C2: 5-hour sitting (interrupted sitting per 30min with 3min light-intensity bodyweight resistance exercise)<br>Duration: 3min/bout x 8min = 24min | Attention (Numeric vigilance test and discrete simple reaction time)<br>Memory (Probed memory test) |
| Wanders et al., 2020 | Crossover RCT: 4 conditions | N = 24 (19 female/5 male)<br>Age: 59.6 ± 8.1 years<br>BMI: 30.2 ± 2.5 kg/m<sup>2</sup> (overweight/obese)<br>Cognitive status: healthy | C1 (HPLF) & C3 (West): 4-hour sitting | C2 (HPLF) & C4 (west): 4-hour sitting (interrupted sitting per 30min with 5min moderate-intensity cycle).<br>Duration: 5min/bout x 6 bouts = 30min | Attention and Executive function (The Test of Attentional Performance Battery) |

Notes. RCT = randomized controlled trial; BMI = body mass index; C=Condition of within-subject crossover design; high mental activity: making puzzles with the CogPack training software; HPLF = high-protein; Low mental activity: watching an informative non-arousing television program called “Rail Away”; low-fat breakfast; West = high in fats/simple sugar & lower in protein/fiber breakfast

## Quality Assessment

The quality assessment of included studies was conducted via the PEDro scale [43]. Of these 13 studies, 10 studies were rated as have a good methodological quality while 3 studies were rated as have only a fair methodological quality. Overall, the methodological quality of the included studies ranged between5 to 8 points. In the majority of the studies the differences of methodological ratings arose from an insufficient reporting of details concerning allocation concealment. In this context, only one study reported subjects blinding. A detailed overview of the methodological rating for each study can be found in Table 2.

**Table 2.** Methodological rating for each study

| Authors | A | B | C | D | E | F | G | H | I | J | K | Total score |
| --- | --- | --- | --- | --- | --- | --- | --- | --- | --- | --- | --- | --- |
| Christmas et al., 2019 | M | 1 | 0 | 1 | 0 | 0 | 0 | 1 | 1 | 1 | 1 | 6 |
| Maasakkers et al., 2020 | M | 1 | 0 | 1 | 0 | 0 | 0 | 1 | 1 | 1 | 1 | 6 |
| Wennberg et al., 2016 | M | 1 | 1 | 1 | 0 | 0 | 0 | 1 | 1 | 1 | 1 | 7 |
| Sperlich et al., 2018 | M | 1 | 0 | 1 | 0 | 0 | 0 | 1 | 1 | 1 | 1 | 6 |
| Wanders et al., 2020 | M | 1 | 0 | 1 | 0 | 0 | 0 | 1 | 1 | 1 | 1 | 6 |
| Block et al., 2018 | M | 1 | 0 | 0 | 0 | 0 | 0 | 1 | 1 | 1 | 1 | 5 |
| Stoner et al., 2019 | M | 1 | 1 | 1 | 0 | 0 | 0 | 1 | 1 | 1 | 1 | 7 |
| Wheeler et al., 2019 | M | 1 | 1 | 1 | 1 | 0 | 0 | 1 | 1 | 1 | 1 | 8 |
| Vincent et al., 2018 | M | 1 | 1 | 1 | 0 | 0 | 0 | 1 | 1 | 1 | 1 | 7 |
| Duvivier et al., 2017 | M | 1 | 0 | 0 | 0 | 0 | 0 | 1 | 1 | 1 | 1 | 5 |
| Charlett et al., 2020 | M | 1 | 0 | 1 | 0 | 0 | 0 | 1 | 1 | 1 | 1 | 6 |
| Bergouignan et al., 2016 | M | 1 | 0 | 1 | 0 | 0 | 0 | 1 | 1 | 1 | 1 | 6 |
| Mullane et al., 2016 | M | 1 | 0 | 0 | 0 | 0 | 0 | 1 | 1 | 1 | 1 | 5 |

*Note:* M=Meet the standards; Total score=4-5= Fair quality; 6-8=Good quality; 9-10=High quality;
A: Eligibility criteria were specified
B: Randomly allocated
C: Allocation was concealed
D: The conditions were similar at baseline
E: Blinding of all subjects
F: Blinding of all therapists
G: Blinding of all assessors
H: Measures of more than 85% of the subjects initially allocated
I: Intention-to-treat analysis

### Meta-analysis

#### Primary outcomes

As listed in the following, significant effects of PE break on cognitive performance and its subdomains (executive function, attention and memory function) with the random-effects model has not been observed during PS: 1): overall cognitive performance: (Hedge g = -0.04, CI 95% -0.16 to 0.07, p = 0.47, I^2^=0.00, p=1.00); 2): memory function: (Hedge g = -0.17, 95% CI -0.70 to 0.36, p = 0.53, I^2^=14.93, p=0.28); 3): executive function: (Hedge g = 0.01,95% CI -0.14 to 0.16, p = 0.90, I^2^=0.00, p=1.00); 4): attention: (Hedge g = -0.11, 95% CI -0.31 to 0.09, p = 0.27, I^2^=0.00, p=1.00). The effects of PE on overall cognition and subdomains were not characterized by heterogeneity.

#### Secondary outcomes

With respect to meta-regression, cognitive performance was not associated with continuous variables: 1) age: β=0.002, 95% CI -0.0077 to 0.0118, p = 0.68; 2) BMI: β= -0.0133, 95% CI -0.0694 to 0.0427, p =0.64; 3) accumulating PE break duration: β= 0.0105, 95% CI -0.1009 to 0.0118, p = 0.85; 4) single PE break duration: β= 0.0029, 95% CI -0.0145 to 0.0202, p = 0.74.

With respect to PE break intensity for cognitive performance, the effects of PE break intervention on cognitive performance was not significantly moderated by PE break intensity (Hedge g = -0.043, CI 95%-0.16 to 0.07, p = 0.47): 1) low PA break [Hedge g = -0.05, CI 95% -0.24 to 0.14, p = 0.58]; 2) moderate PA break [Hedge g = -0.02, CI 95% -0.18 to 0.04, p = 0.82]; 3) high PA break [hedge g = -0.11, CI 95 -0.46 to 0.23, p = 0.52].

#### Publication bias

There was no indication of publication bias for cognitive function as indicated by funnel plots. Egger’s tests suported that there is no funnel plot asymmetry: (Egger’s P=0.873),

## Discussion

Accumulating evidence has demonstrated that acute (single) bouts of PE can exert a positive effect on cognitive performance across a wide range of populations [24-27]. However, so far, no systematic review or meta-analysis has focused on the effect of breaking up PS by PE breaks on cognitive performance. Given the rising prevalence of physical inactivity in general and PS (as part of sedentary behavior) in particular across industrialized nations, such effects have direct implications for public health. To address this gap in the literature, this systematic review and meta-analysis aimed to answer the following main research questions: (i) Does breaking up PS by acute PE breaks lead to improvements concerning measures of cognitive performance? and (ii) Does specific exercise characteristics of the PE breaks (e.g., type of physical exercise, exercise intensity, and/or exercise duration) moderate the effect of PE breaks during PS on cognitive performance?

### Main study findings

Although some single studies reported some significant improvements on executive function, memory, and composite scores of cognitive function when PE break was compared to PS condition, our meta-analysis did not provide any evidence for a significant effect of acute PE breaks during PS on cognitive performance as compared to PS. Moreover, we did not find evidence for a possible dose-response relation concerning exercise characteristics of acute PE breaks and changes in measures of cognitive performance.

### Possible explanation for study findings

The majority of the reviewed studies had a relatively small sample size and our meta-analysis was based on nine studies, meaning that our findings should be interpreted with caution. This unexpected finding could be caused by a myriad of factors (e.g., methodological differences). Among those factors, the relatively large heterogeneity concerning exercise characteristics of the acute PE break as well as the control condition could be important confounders. With regard to the exercise characteristics, there is evidence that exercise variables such as exercise intensity influence the effects of acute physical exercises on cognitive performance [24, 27, 53, 54].

With respect to exercise intensity and exercise duration of the acute PE breaks, it was reported that in older adults 2-min light-intensity PE break[16] or 5-min moderate-intensity PE break[49] following 30-min sitting did not benefit measures of cognitive performance (i.e., attention and executive function), while Wheeler and colleagues found that interrupting PS with a bout of 30-min moderate intensity walking induce positive effects on measures of cognitive performance (i.e., executive function) [51]. As those mixed evidence does not allow to draw solid conclusions, further high-quality studies are needed to rule out possible dose-response relationships concerning exercise intensity and acute change in cognitive performance. In this context, it should be considered that in sedentary and inactive older adults, PE breaks with a light exercise intensity might be too weak to simulate significant cognitive improvements immediately after the acute PE breaks[50] although there are undoubtedly several other variables influencing this relationships (e.g., initial fitness level of the participant, delay between acute exercise cessation and cognitive testing). Thus, in this age group exercises at moderate or high exercise intensity might be better situated for acute PE breaks aiming to positively influence cognitive performance. However, given the inconsistent findings in available studies either showing or showing no benefit of acute PE breaks, further trials are necessary to provide further support for these assumptions.

Concerning younger adults, either interrupting PS by 3 min of light-intensity walk or bodyweight resistance exercises could not improve cognitive performance [45, 47], whereas 3-min moderate walk has a positive effect on executive functioning [36]. In children, interrupting PS by 2-min of low-, moderate-, or high-intensity exercise every 20 minutes was not effective in promoting cognitive benefits concerning executive functioning [52]. Taken together, available evidence concerning the influence of exercise intensity of acute PE breaks is rather mixed, and thus future studies investigating possible dose-response relationships of exercise intensity and cognitive performance are needed to draw more robust conclusions.

With respect to exercise frequency, most of the included studies chose to break up PS by PA break every 30 minutes, but only two studies [36, 51] found significant improvements concerning executive functioning. Speculatively, interrupting PS every 30 minutes might be too frequent as there is evidence from educational psychology suggesting that the concentration level increases and reach a maximum at 10-15 min in class. Thereafter, the concentration gradually decreases to baseline levels (achieved around 50 minutes in class), although, the level of concentration at 30 minutes in class is still higher than baseline [55]. Following this line of interpretation, acute PE breaks every 30 minutes may impair concentration and/or attention rather than leading to cognitive improvements. In line with this assumption, Mullane et al. observed in younger adults’ cognitive improvements in response to acute PE breaks being conducted every hour [37]. In order to get a more comprehensive understanding whether the frequency of PA break moderate possible effects of acute PE breaks on cognitive performance, further studies with different frequencies of PE breaks are urgently needed.

There is some evidence in the literature that the type of physical exercise can also have an considerable impact on the effectiveness of acute physical exercises interventions (e.g., acute PE breaks) on measures of cognitive performance [27]. In our systematic review and meta-analysis, three studies of the included studies [36, 37, 51] observed positive effects on cognitive performance when standing, walking and/or cycling were used. For example, Mullane et al. noticed that cycling improved cognitive performance to a greater extent than standing or walking. However, as the study of Mullane and colleagues was the only study that directly compared the effect of different types of physical exercises, no generalizable conclusion can be drawn. Thus, further research is needed to determine whether different types of physical exercises used during acute PE breaks during PS might influence cognitive performance differentially.

In addition to exercise characteristics, also the choice of the control condition might have influenced the findings. For instance, there is evidence in the literature that different activities being entailed in the definition of sedentary behavior are differentially associated with cognitive performance. For instance, sitting and watching TV is negatively associated with cognitive performance while computer-use (also performed in a sitting position) was positively associated with cognitive performance [13]. Thus, the degree of mental stimulation being allowed or provided in the control condition might also influence the current findings.

Moreover, based on the Canadian 24-hour Movement (24-HMB) Guidelines for Children and Youth, a greater attention should be paid on an integrated and holistic perspective including factors such as sleep, sedentary behavior and physical activity rather than look upon single factors (e.g., only physical activity) [56]. Accordingly, assessing of 24-HMB might be of relevance for studies investigating the effects of acute PE breaks during PS on cognitive performance as, there is increasing evidence that healthy sleep patterns are associated with favorable outcomes concerning the performance in most domains of cognitive functioning [57]. For instance, sleep deprivation is associated with lower cognitive performance [58]. Hence, participants should be advised to maintain adequate sleep to avoid a possible diminishing of the effect of PE break interventions on cognitive performance that can be caused by inadequate sleep patterns. With respect to the studies included in the current systematic review and meta-analysis, only five assessed sleep behaviors. Exemplarily, Vincent et al. investigated the effect of breaking PS by PA on cognitive function under different conditions of sleep restriction and noticed that only 5-h sleep may negatively influence cognitive performance, which, in turn, suggested that an adequate sleep duration (e.g., 8-9 hours’ sleep opportunities) are necessary. Future studies should aim to more rigorously control for sleep by using objective measures (e.g., via Actigraphy) and subjective measures of sleep (e.g., sleep quality via Pittsburgh Sleep Quality Index).

Although we did not find compelling evidence that acute PE break during PS have a positive effect on measures of cognitive performance, we hold the opinion that breaking up PS by PE is a valuable strategy being worth to be implemented in our daily lifes as it can reduce our feelings of fatigue, sleepiness and increase the levels of self-perceived energy and vigor, relieves feeling of physical discomfort. For example, Wennberg et al. observed that in older overweight/obese adults, compared to keeping prolonged sitting for 7 hours, interrupted sitting per 30 min with 3min light-intensity walk can reduce fatigue levels at 4h and 7h [50]. In a comparable manner, Vincent et al. reported that participants who engaged in 8-hour sitting (SIT) or interrupted sitting per 30min with 3min light-intensity walk (ACT) reported higher levels of sleepiness, and felt less alert in the SIT condition compared with the ACT condition [47]. These findings suggest that adults who maintain a sedentary lifestyle can benefit from acute PE breaks during PS when psycho-emotional parameters are considered although such changes those not readily transfer to cognitive performance improvements.

### Strength and limitations

A strength of this meta-analysis is, that, as compared to previous meta-analytical reviews investigating the effects of PE on cognitive function, our research team firstly summarized the existing literature on the effects of acute PE breaks on cognitive measures during PS. Furthermore, given that SB has detrimental effects on physical and mental (cognitive) health, this emerging research field is from great public interest. In this context, in the past 30 years the rational educational model to promote regular PE engagement has been relatively unsuccessful, thus breaking up PS by acute PE breaks may be another readily acceptable way to get individuals actively at the workplace and support the importance to enlarge our knowledge in this direction.

There are some limitations that need to be acknowledged. Firstly, studies that used standing as the acute PE break alone or investigated solely the influences of acute PE breaks on brain activities were excluded. Secondly, only studies on a total PS duration of ≧ hours were included, which may exclude other relevant studies.

## Conclusion

The findings of the current systematic review and meta-analysis suggest that breaking up PS by acute PE breaks does not improve, but also not impair cognitive performance. Consequently, individuals may use physical activity breaks to promote their physical health without compromising their cognitive performance. This finding has a high practical relevance for the work-life in many countries as there is an increasing tendency towards work performed while standing or sitting. Given that our finding was informed by a limited number of studies, a re-assessment is encouraged to validate the lack of effects on cognitive performance.

## Data Availability

All data produced in the present study are available upon reasonable request to the authors

## Data Availability Statement

All data produced in the present study are available upon reasonable request to the authors.

